# Tibiofemoral Contact Loads across Walking, Kneeling, and Jumping Tasks with and without Cognitive Challenges

**DOI:** 10.1101/2025.08.16.25333778

**Authors:** Scott M. Monfort, Devon Doud, Fatemeh Aflatounian, Corey Pew, David J. Saxby, David F. Graham

## Abstract

**Background:** Characterizing knee contact loads across diverse movements and in the presence of cognitive challenges is necessary to understand occupational factors associated with knee osteoarthritis risk in manual laborers (e.g., agricultural workers).

**Research Questions:** What are the tibiofemoral contact forces during walking, kneel-to-stand, and jump landing and how do cognitive challenges affect these contact forces?

**Methods:** Twenty-four healthy manual laborers and recreational athletes completed walking (WALK), standing from a kneeling position (KNEEL), and jump landings (JUMP), each with a cognitive challenge (COG) and without (BASE). Neuromusculoskeletal modelling was used to estimate tibiofemoral contact forces. Peak tibiofemoral contact forces (medial, lateral, and total) during the stance phase were primary outcomes. Correlations between peak contact forces across tasks and comparisons of contact force time series were also evaluated.

**Results:** There were no significant Task*Condition interactions or main effects of Condition; however, there was a main effect of Task for peak medial, lateral, and total tibiofemoral contact forces (*P*<0.001). Tibiofemoral contact forces during WALK were poorly related to corresponding estimates during KNEEL or JUMP (r=0.13±0.08), but KNEEL and JUMP were significantly related (r=0.64±0.16). COG resulted in increased medial contact forces during the 28-66% of stance phase for KNEEL.

**Significance:** The differences in loading magnitude alongside a lack of rank-order consistency between contact loads during walking with those during more dynamic tasks motivates the need to sample a representative range of motor tasks and situational demands for the population of interest to reflect real-world contact loads and study their relation to knee osteoarthritis.

**Highlights:**

- A reactive challenge increased medial knee loading when rising from kneeling
- Cognitive challenges had variable and often negligible effects on contact forces
- Rising from kneeling had increased peak knee contact forces than walking
- Peak tibiofemoral contact forces for rising from kneeling were at deep knee flexion
- Knee contact forces during walking did not relate to those during other tasks

## Introduction

Deep knee flexion tasks result in larger knee contact forces than in walking [1–3] and have been associated with increased risk of knee osteoarthritis (OA) [4–6]. Prior studies on tibiofemoral loading during deep knee flexion tasks have largely focused on variations of squatting [2, 3] or stair climbing [7, 8], with few investigations into rising from a kneeling position and limited to a sagittal plane knee model [1]. Therefore, insight into medial-lateral load distribution and comparison of these contact forces across motor tasks may aid us to better understand the factors contributing to disproportionate knee OA risk in occupations that commonly perform deep knee flexion movements, such as manual laborers (e.g., agricultural workers) [9–11].

The effects of cognitive challenges on knee contact forces during real-world tasks have not been investigated. Dual-tasks (e.g., concurrently performing a motor and cognitive task) can reduce stability, decrease movement speed, and increase muscle activation [12–15]. Notably, dual-task conditions may better reflect movement patterns of daily life compared to laboratory evaluation of isolated motor function [16]. However, the effects of dual-task conditions on tibiofemoral contact forces across a range of movements remain unexamined.

Neuromusculoskeletal computational modeling enable estimation of joint contact forces [17, 18]. Computational approaches that use individuals’ muscle activation patterns to inform the modeled muscle forces are well aligned to account for the potential effects of cognitive challenges task-specific muscle activation patterns [19]. Specifically, an electromyography (EMG)-informed modeling approach enables investigation into the effect of different movements and cognitive conditions on tibiofemoral contact forces while accounting for task-, condition-, and individual-specific muscle activation patterns.

To better understand contact forces during deep flexion tasks and the potential for cognitive challenges to influence tibiofemoral contact forces, the purpose of this study was to model tibiofemoral contact forces during walking, kneel-to-stand, and jump landing tasks during single– and dual-task conditions. In addition, we hypothesized that adding a cognitive challenge would result in increased contact forces.

## Methods

Manual laborers and recreational athletes were recruited for the study. To be eligible, they were required to be physically active, healthy adults between 20 and 60 years old with a Western Ontario and McMaster Universities Osteoarthritis Index pain score less than two [20], no history of lower body surgery, no lower body injury in the past three years, and a Tegner activity scale greater than three. The study protocol was approved by the University’s Institutional Review Board, and written informed consent was obtained from all participants.

### Biomechanical Testing

Motion capture testing was performed as participants completed walking (WALK), standing from a kneeling position (KNEEL), and jumping landings (JUMP) both with (COG) and without (BASE) the addition of a cognitive challenge. For WALK, participants were instructed to walk normally through the laboratory while staring at a fixation cross (BASE) or completing a visual Stroop test (COG) [21]. Three good trials (i.e., clean force plate contact, speed within 10% of self-selected pace) were collected.

The BASE KNEEL task required participants to kneel with one knee on a force plate and the foot of the non-kneeling limb resting on a separate force plate. Participants then stood as quickly as they could without having their arms contact their lower body. For the COG condition, participants stood when a predesignated “Go” color was displayed by one of two peripheral FITLIGHT sensors (Aurora, Ontario, Canada). Stance phase was defined as contralateral knee liftoff (<20N) to contralateral foot strike (>20N). Three good trials (i.e., compliant with task instructions) were collected.

The JUMP task consisted of participants jumping off of a 30-cm box over a distance of half of their height to land on force plates [22]. As participants landed, they were instructed to immediately complete a maximal effort jump. For the BASE condition of the task, participants jumped straight up for this secondary jump. For the COG condition, a directional cue was displayed on a screen in front of them ∼250 msec prior to the initial landing. The cue indicated whether the secondary jump should be straight up, or 45° to the right or left. Only the initial landing for trials with a secondary jump direction of straight up were analyzed. Three good trials (i.e., clean force plate contact, compliant with task instructions) were collected.

Ten motion capture cameras (Motion Analysis Corporation, Rohnert Park, CA, USA) collected data at 250 Hz and five force plates (AMTI, Watertown, MA, USA) collected at 1,000 Hz. Additionally, eight surface electromyography sensors (Delsys Trigno, Natick, MA, USA) collected data at 1,000 Hz from eight muscles of the dominant limb (gluteus maximus, gluteus medius, rectus femoris, vastus lateralis, semimembranosus, bicep femoris, tibialis anterior, and medial head of gastrocnemius) following SENIAM guidelines (seniam.org). A markerset by Graham et al. was used [23]. In brief, it included tracking clusters on the thighs and shanks and markers on the metatarsals, calcanei, malleoli, femoral epicondyles, anterior and posterior superior iliac spines, and iliac crests. The upper body had a modified Plug-in Gait markerset.

### Data Preparation

Data were prepared for neuromusculoskeletal modeling using MOtoNMS [24]. Marker and force data were filtered with a 2^nd^-order lowpass Butterworth filter with cutoff frequencies of 6 Hz (WALK, KNEEL) and 15 Hz (JUMP). EMG data were bandpass filtered (30-500 Hz), full wave rectified, and then lowpass filtered with a 6 Hz cutoff frequency [18]. The resulting linear envelopes for each muscle were scaled to muscle-specific maximums of the linear envelopes across trials for a given participant. EMG quality for each trial was evaluated using a validated deep neural network tool [25]. EMG signals for a given trial that were characterized as ‘Noise’ were replaced with synthesized signal during neuromusculoskeletal modeling, as were muscle-tendon units (MTUs) that did not have experimental EMG data.

### Musculoskeletal Model

OpenSim v4.1 [26] was used via MATLAB v2023b (MathWorks, Natick MA, USA). The Catelli 37-degree of freedom musculoskeletal model with 80 Hill-type muscle tendon units (MTU) [27] was adapted for this study. Humerus, ulna, radius, and hand bodies and their associated 14 degrees of freedom were omitted due to occasional poor tracking of sparsely markered arms during tasks with large arm motion (i.e., JUMP). The model was further augmented to have planar hinge knees to enable compartmental tibiofemoral contact force estimation (see **Supplemental Material** for additional details) [28].

The generic model was linearly scaled using the OpenSim Scaling Tool. The optimal fiber length and tendon slack length MTU parameters were then further refined to retain dimensionless operating ranges over the scaled model range of motion [29]. Finally, maximum isometric force parameters were adjusted based on participants’ height and mass [30].

### Neuromusculoskeletal Modeling

Inverse kinematics, inverse dynamics, and muscle analyses were performed using OpenSim v4.1. Calibrated EMG-informed neuromusculoskeletal modeling (CEINMS) was then used to estimate muscle contributions to the observed biomechanics and as the basis for estimating tibiofemoral contact forces [17, 31]. The eight EMG signals were mapped to 22 MTU excitations for the instrumented limb, with synthesized signals used for MTUs without EMG data (unmeasured or unsuitable data quality) [19, 32]. CEINMS calibration (v0.21.1) was performed using a subset of trials (1-2 of each movement task) for each participant to further optimize MTU parameters. A contact force term was included in the calibration objective function to align with prior research [28], and because prior work suggests it may obtain more physiologically plausible contact force estimates [17, 33].

The CEINMS calibrated model was then used for CEINMS execution (v0.21.0) in “EMG-assisted” mode to estimate MTU excitations for trials not used in the calibration step [31]. Outputs of the CEINMS execution step were evaluated to determine their ability to generate joint torques that tracked hip, knee, and ankle sagittal plane net joint moments estimated through inverse dynamics (>80% R^2^ and <7% normalized root mean squared error). A summary of final CEINMS calibration and execution parameters along with additional detail on the calibration process is provided in the **Supplemental Material**.

Medial and lateral tibiofemoral contact forces (MTF and LTF, respectively) as well as total tibiofemoral contact forces (TTF=MTF+LTF) were estimated using a point contact approach with the planar hinge knee model [34]. Peak contact forces (MTF, LTF and TTF) during stance phase were the primary outcomes, with the time series of the contact forces throughout the stance phases used to provide additional insight.

### Statistical Analysis

Statistical analysis was performed using R (version 4.4.1) via RStudio (version 2025.5.0 + 496, Posit Software, PBC). The effect of the movement task and condition on the peak TTF, MTF, and LTF were analyzed using linear mixed-effects models. Participant was a random factor, and movement Task (WALK, KNEEL, and JUMP) and Condition (BASE, COG) were fixed factors. Age, BMI, and walking speed (for WALK) were considered as covariates, with models run with and without their inclusion. Significance level was set at α=0.05/3=0.0167 to account for the three primary outcomes (peak TTF, MTF, LTF). When appropriate, post-hoc Tukey pairwise comparisons were made. Additional details on statistical analyses provided in **Supplemental Material**.

To provide further context, two additional analyses were included. First, Spearman correlations between peak contact forces across movement Tasks were used to determine the extent that the ranking of contact forces remained consistent across tasks. Finally, paired T-test statistical parametric mapping analyses were performed using SPM1D (version M.0.4.12, www.spm1d.org) to compare the tibiofemoral contact forces for BASE and COG conditions, with the movement tasks considered independently. This analysis was done to identify altered loading patterns that may exist throughout stance phase.

### Sample Size Justification

A sample size of n=13 was determined to provide 92% statistical power to detect a main effect of Condition at α=0.0167. Additional details are available in **Supplemental Material**.

## Results

Twenty-four participants (25.6±5.4 years; 21 male/ 3 female; 1.77±0.09 m; 73.4±11.1 kg) had viable EMG data quality to be included in the current study. Data for two participants were omitted for JUMP and one participant was omitted for KNEEL due to the CEINMS moments not meeting the thresholds for agreement with inverse dynamics moments. The final dataset consisted of 82% out of the targeted 432 trials (24 participants x 3 tasks x 2 conditions x 3 trials), with the median number of trials averaged for each participant-task-condition being 2-3 (see **Supplemental Material** for additional quality metrics). The average of the root mean square marker error for inverse kinematics was 1.6±0.5 cm, meeting recommended guidelines [35].

Total contact force estimates during WALK (Total: 3.6±0.7 Bodyweights [BW]) were slightly higher than a study implementing a similar modeling approach (Total: 2.8±0.6 BW) [28], albeit well within two standard deviation agreement recommendation [35].

There were no significant Task*Condition interactions or main effects of Condition for TTF, MTF, or LTF (**Table 1**), although an interaction for the MTF trended towards significance (*P*=0.032) (**Figure 1**). A follow-up analysis indicated that the trending interaction was driven by WALK having decreased contact forces during COG compared to BASE (*P*=0.006), while KNEEL and JUMP had nonsignificant increases in MTF during COG relative to BASE (*P*>0.05). The change in MTF was no longer significant (*P*=0.29) when including walking speed as a covariate, suggesting that the decrease in walking speed from BASE to COG (1.37±0.17 m/sec to 1.27±0.15 m/sec, respectively) explained the decrease in MTF.

**Figure 1.**
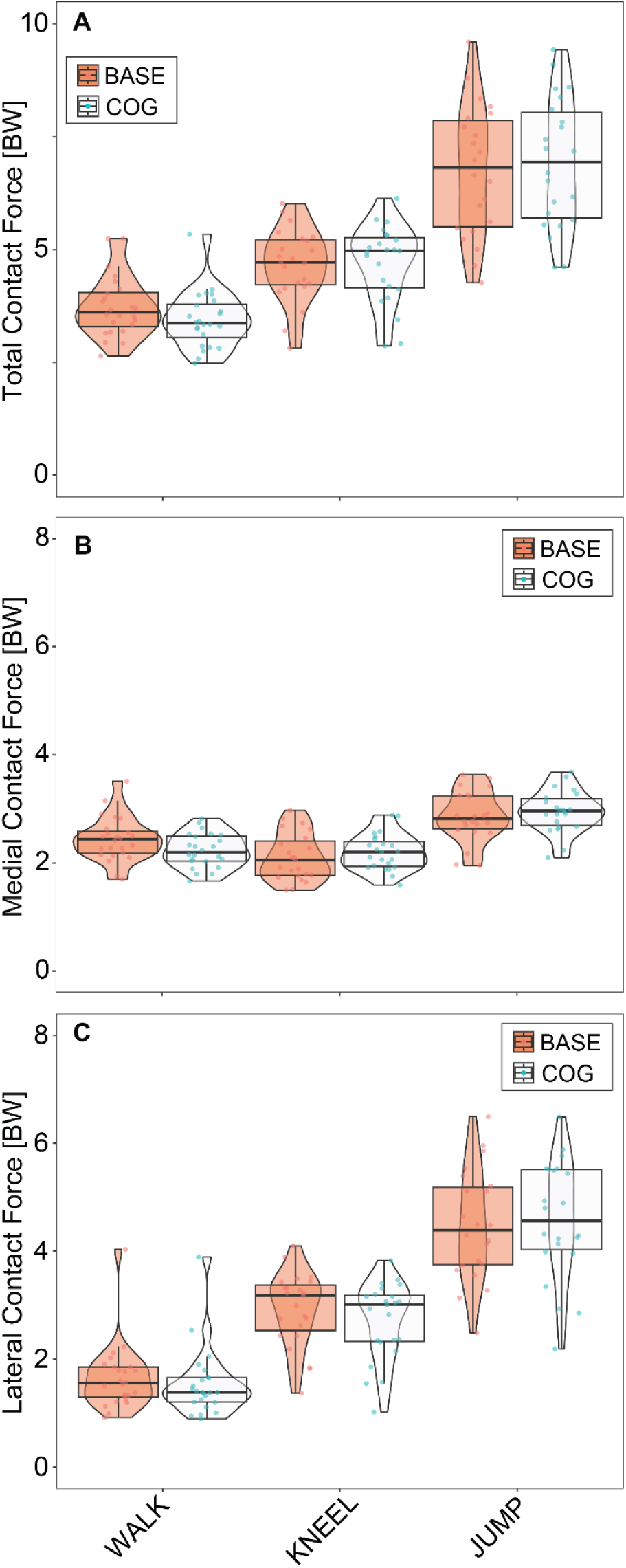
Peak total (A) medial (B) and lateral (C) tibiofemoral contact forces across tasks and conditions.

**Table 1.** Summary of mixed effects models. P-values are presented.

|  | **Task** | **Condition** | **Task\*Condition** |
| --- | --- | --- | --- |
| **Total** | <0.0001* | 0.81 | 0.34 |
| **Medial** | <0.0001* | 0.92 | 0.032 |
| **Lateral** | <0.0001* | 0.49 | 0.45 |

\* Indicates statistical significance ( $p < 0.0167$ )

There was a main effect of Task for TTF, MTF, and LTF (all p<0.001, **Figure 1**). JUMP produced the largest MTF (95% CI: 2.75-3.02 BW), LTF (95% CI: 4.23-4.80 BW), and TTF (95% CI: 6.45-7.17 BW). Contact forces during KNEEL were higher than WALK for TTF (95% CI: 4.31-5.02 BW vs. 3.23-3.94 BW) and LTF (95% CI: 2.53-3.10 BW vs. 1.32-1.87 BW); however, MTF was higher for WALK (95%CI: 2.20-2.47 BW) than KNEEL (95% CI: 2.01-2.28 BW).

The correlation analysis indicated that tibiofemoral contact forces during WALK were poorly related to corresponding estimates during KNEEL or JUMP (i.e., not significant and r=0.13±0.08), but that KNEEL and JUMP were significantly related (r=0.64±0.16) (**Figure 2**). Furthermore, BASE and COG estimates were significantly related for WALK (average r=0.71), KNEEL (average r=0.83), and JUMP (average r=0.89). Similar correlations were observed for MTF and LTF **(Supplemental Figure 1**).

**Figure 2.**
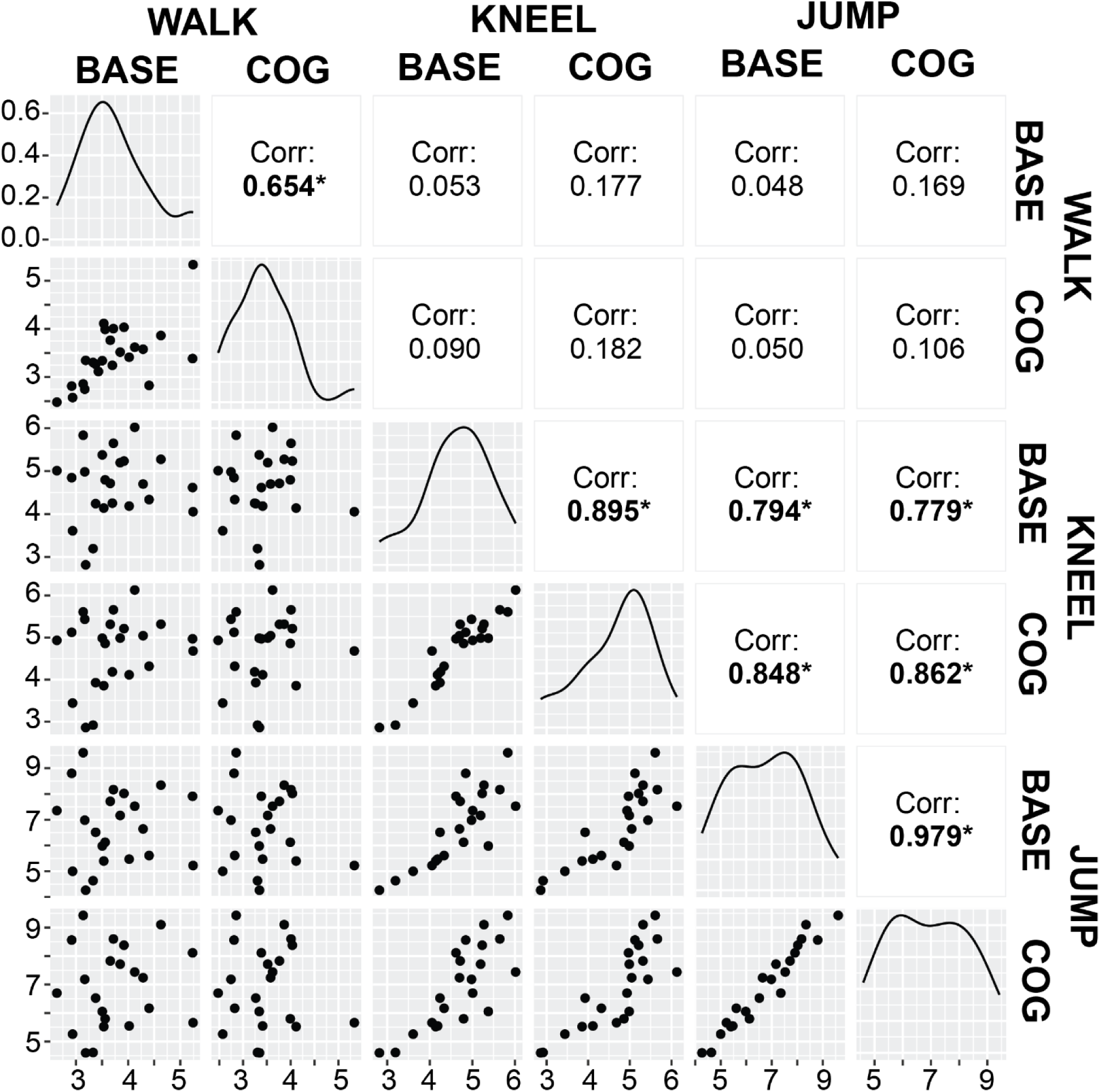
Correlations for total tibiofemoral contact forces across combinations of tasks and condition. Spearman correlation coefficients are reported in the upper right portion of the figure, with * indicating statistical significance. Lower left panels reflect the scatterplots of total tibiofemoral contact force with units of BW for the respective task-condition combination. The diagonal panels display the distribution of the variable.

The time series analysis corroborated the limited effect of Condition on contact loads; however, differences between BASE and COG were seen at select portions of the KNEEL task (TTF: 0-3% stance phase; MTF: 28-66%; LTF: 0-12%; **Figure 3**). Differences were more prevalent for rebalancing the contact force between medial and lateral compartments rather than changing the total tibiofemoral contact force. Similar time series were observed between BASE and COG for WALK and JUMP, with differences limited to a brief increase (9-12% stance phase) in total contact force for BASE relative to COG for WALK (**Supplemental Figure 2**).

**Figure 3.**
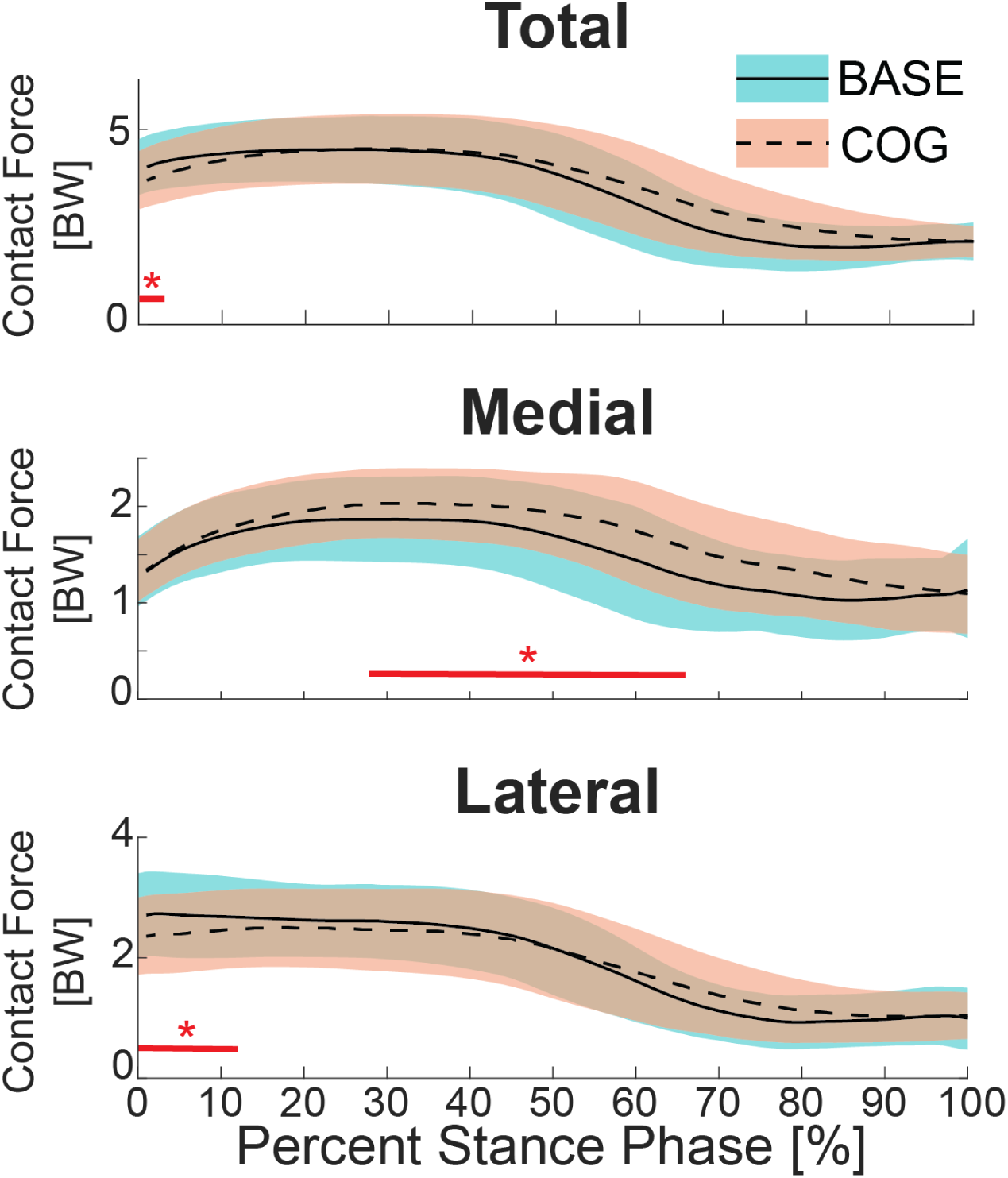
Time series of total, medial, and lateral tibiofemoral contact forces throughout the KNEEL task. Red lines with an asterisk along the x-axis indicate regions that are statistically different based on paired T-test SPM analyses.

## Discussion

In this study, we characterized the tibiofemoral contact forces across a range of tasks to test the hypothesis that performing these tasks with added cognitive challenge would increase tibiofemoral contact force. Total magnitude and medial-lateral distribution of tibiofemoral contact forces differed between walking, kneel-to-stand, and jumping tasks. Our hypothesis was largely refuted with minor differences in contact forces due to added cognitive challenge, although the nature of the cognitive challenge (e.g., reactive vs. working memory) influenced these effects. Additionally, contact forces during walking (single-or dual-task) were poorly related to contact forces during more demanding tasks of kneel-to-stand and jumping, suggesting gait biomechanics are likely insufficient to represent the biomechanical loading for manual laborers who routinely perform more demanding physical tasks. Sampling a representative range of motor tasks and situational demands for the population of interest is needed to reflect real-world contact loads and study their relation to knee OA.

Differences in tibiofemoral contact forces were observed across tasks, with TTF increasing with increases in task intensity (i.e., WALK<KNEEL<JUMP). Notably, the KNEEL task resulted in one additional bodyweight of tibiofemoral contact force relative to walking. TTF during KNEEL were ∼1.6 BW lower on average than reported by Nagura et al., which may reflect our inclusion of a contact force term when calibrating the CEINMS model that has shown prior benefit in attaining physiologically plausible contact forces [17, 33]. When compared across studies, our KNEEL contact forces were higher than prior modeling estimates of sit-to-stand tibiofemoral contact forces [36], generally higher than estimates during stair climbing [7, 8], and higher or similar to deep squatting [2, 3], although some studies report higher values [1, 37].

Our findings suggest KNEEL decreases the medial compartment load sharing compared to WALK, which has been reported for stair ascent and descent with older adults with and without OA of varying severity [7, 8]. A post-hoc investigation into medial load share (i.e., MTF divided by TTF) corroborated this observation. Medial load share was below 50% at the instant of peak TTF for KNEEL (43%±8%) and JUMP (38%±7%) but not WALK (63%±12%). Our study extends this consideration to a kneeling task with deep knee flexion angles (maximum knee flexion: 121°±6°). In addition to the lateral bias, the peak contact forces occurred at deep knee flexion, with greater knee flexion for KNEEL (106°±12°) compared to JUMP (72±13°). Higher contact forces occurring at increased knee flexion poses a potential risk for an adverse cartilage response due to the decreased tibiofemoral contact area in deeper knee flexion causing similar contact forces to result in relative increases in stresses experienced by cartilage [38].

Notably, moderate-to-strong correlations were observed between KNEEL and JUMP loading, suggesting the tendency for individuals to perform these tasks with greater or lesser contact forces was consistent. This association was in contrast to WALK, where tibiofemoral contact forces did not associate with those during KNEEL or JUMP. The lack of association of WALK loads to those seen in more dynamic tasks that impose greater cartilage loading suggests that additional tasks are necessary to establish representative estimates of real-world cartilage loading. This need may be increased in manual labor occupations where individuals frequently perform demanding tasks in postures that involve increased hip and knee flexion.

Cognitive challenges had nuanced effects on tibiofemoral contact forces. No significant main or interacting effects involving cognitive condition were observed for peak TTF, MTF, or LTF; however, cognitive challenge dependent effects were observed during the SPM analysis of the entire stance phase. Movement speed may explain the differences in response to cognitive challenges used in this study, where cognitive challenges that elicited faster movement (KNEEL) were associated with isolated increases in contact forces, the standardized jump landing approach retained a consistent approach movement speed between BASE and COG for JUMP and resulted in no differences in contact forces, and the Stroop task resulted in decreased walking speed and isolated decreases in contact forces. Therefore, the potential for cognitive challenges to alter overall task performance (e.g., movement speed) is a potentially salient mechanism in which real-world challenges may alter tibiofemoral contact forces within a task.

There are some limitations that should be considered when interpreting the findings from this study. First, the tibiofemoral contact forces are estimates from neuromusculoskeletal modeling rather than *in vivo* measurements. However, the use of a model suitable for deep flexion tasks, an established tibiofemoral contact force estimation technique, and EMG-informed neural mode to solve the muscle redundancy problem that was calibrated with the inclusion of a term to penalize high tibiofemoral contact forces are supported by the literature and provide a well-justified approach for obtaining plausible contact forces [17]. Secondly, because a limited subset of movement and cognitive tasks were included in the study, the limited effects of cognitive challenges on tibiofemoral contact forces found in this study should not be overgeneralized.

## Conclusion

Total knee contact loads ranged from 3.6-6.8 BW across three tasks that reflect a range of movements potentially encountered during manual labor. A reactive cognitive challenge elicited increased medial tibiofemoral contact force during a portion of stance when rising from kneeling; however, other distracting and reactive cognitive challenges did not significantly influence tibiofemoral contact forces. The lack of rank-order consistency between contact loads during walking with those during more dynamic tasks motivates the need to sample a representative range of motor tasks and situational demands for the population of interest to reflect real-world contact loads and study their relation to knee OA.

## Supporting information

GLIMMPSE Power Analysis Settings File

## Data Availability

All data produced in the present study are available upon reasonable request to the authors

