## Supplemental Material for "Tibiofemoral Contact Loads across Walking, Kneeling, and Jumping Tasks with and without Cognitive Challenges"

***Methods - Modifications to Musculoskeletal Model:***

The Catelli 37-degree of freedom musculoskeletal model with 80 Hill-type muscle tendon units (MTU) [1] was adapted for this study. Humerus, ulna, radius, and hand bodies and their associated 14 degrees of freedom were omitted due to occasional poor tracking of sparsely markered arms during tasks with large arm motion (i.e., JUMP).

The model was augmented to have planar hinge knees to enable compartmental tibiofemoral contact force estimation [2]. Specifically, a sagittal knee body and an auxiliary intercondylar body were added to each knee limb that were used to create medial and lateral knee hinge joints fixed in the tibia body. The intercondylar bodies of the generic template model were set at 0.04 meters and then scaled based on the distance between markers on the medial and lateral femoral epicondyles during the OpenSim scaling step. Although other models have been developed to allow for compartmental tibiofemoral forces during deep flexion tasks [3], we used the planar knee approach because it allowed for compartmental tibiofemoral contact forces to be incorporated into the objective function during neuromusculoskeletal modeling.

***Additional Details on Neuromusculoskeletal Modeling Workflow:***

*CEINMS Calibration*

CEINMS calibration (v0.21.1) was performed using a subset of trials (1-2 of each movement task) for each participant to further optimize MTU parameters, within the bounds of 0.5-1.5, 0.9-1.1, and 0.9-1.1 of the uncalibrated model values for model strength, tendon slack length, and optimal fiber length, respectively. The calibration objective function included terms to: 1) minimize error in torques (relative to inverse dynamics), 2) minimize the maximum sum of peak excitation across MTUs of the instrumented limb, and 3) minimize tibiofemoral contact forces. The calibration objective function included terms to: 1) minimize error in torques (relative to inverse dynamics), 2) minimize the maximum sum of peak excitation across MTUs of the instrumented limb, and 3) minimize tibiofemoral contact forces. The relative weighting of the contact force term was adjusted to be approximately a third of the torque error term to prioritize the torque tracking during calibration.

*CEINMS Execution*

The CEINMS calibrated model was then used for CEINMS execution (v0.21.0) for trials not used in the calibration step. EMG-assisted mode was used to estimate MTU excitations that minimized the following objective function [4]:

$$\boldsymbol{\alpha} \sum_{d}^{DOFs} (\bar{M}_d - M_d)^2 + \boldsymbol{\beta} \sum_{j}^{MTU_{all}} e_j^2 + \boldsymbol{\gamma} \sum_{k}^{MTU_{emg}} (\bar{e}_k - e_k)^2$$

where $\bar{M}_d$ and $M_d$ are measured and estimated joint moments, respectively; $e_j$ is muscle-tendon unit (MTU) excitation for all MTUs for the instrumented limb; and $\bar{e}_k$ and $e_k$ are experimental and adjusted excitations, respectively, for MTUs having EMG-based excitations being adjusted. α, β, and γ are weighting factors.

Objective function weightings α and β were set at one for all calculations. γ was initially set at 75 and was iteratively decreased to a minimum of one. In the event that torques were unable to meet criteria for agreeing with inverse dynamics torques even with γ=1, the CEINMS calibration step was repeated with a decreased weighting for the contact force term, and then the CEINMS execution step was repeated. A summary of final CEINMS calibration and execution parameters is provided in **Supplemental Table 1**.

**Supplemental Table 1.** Calibration and execution parameter values for each participant.

| Calibration | | | | | | Execution | | |
|---|---|---|---|---|---|---|---|---|
| TorqueErrorNorm | | MuscleForce_MaxSumPeak | | ScalarContactForce | | | | |
| Weight | Exponent | Weight | Exponent | Weight | Exponent | α | β | γ |
| 1 | 1 | 1 | 1 | 1.E-04 | 2 | 1 | 1 | 50 |
| 1 | 1 | 1 | 1 | 1.E-04 | 2 | 1 | 1 | 25 |
| 1 | 1 | 1 | 1 | 2.E-04 | 2 | 1 | 1 | 75 |
| 1 | 1 | 1 | 1 | 5.E-05 | 2 | 1 | 1 | 10 |
| 1 | 1 | 1 | 1 | 1.E-05 | 2 | 1 | 1 | 25 |
| 1 | 1 | 1 | 1 | 1.E-04 | 2 | 1 | 1 | 25 |
| 1 | 1 | 1 | 1 | 1.E-04 | 2 | 1 | 1 | 25 |
| 1 | 1 | 1 | 1 | 1.E-04 | 2 | 1 | 1 | 10 |
| 1 | 1 | 1 | 1 | 1.E-04 | 2 | 1 | 1 | 25 |
| 1 | 1 | 1 | 1 | 1.E-04 | 2 | 1 | 1 | 1 |
| 1 | 1 | 1 | 1 | 1.E-04 | 2 | 1 | 1 | 10 |
| 1 | 1 | 1 | 1 | 1.E-04 | 2 | 1 | 1 | 1 |
| 1 | 1 | 1 | 1 | 1.E-04 | 2 | 1 | 1 | 1 |
| 1 | 1 | 1 | 1 | 1.E-04 | 2 | 1 | 1 | 25 |
| 1 | 1 | 1 | 1 | 1.E-04 | 2 | 1 | 1 | 10 |
| 1 | 1 | 1 | 1 | 5.E-05 | 2 | 1 | 1 | 10 |
| 1 | 1 | 1 | 1 | 1.E-04 | 2 | 1 | 1 | 10 |
| 1 | 1 | 1 | 1 | 2.E-04 | 2 | 1 | 1 | 75 |
| 1 | 1 | 1 | 1 | 1.E-04 | 2 | 1 | 1 | 25 |
| 1 | 1 | 1 | 1 | 1.E-04 | 2 | 1 | 1 | 75 |
| 1 | 1 | 1 | 1 | 5.E-05 | 2 | 1 | 1 | 1 |
| 1 | 1 | 1 | 1 | 1.E-05 | 2 | 1 | 1 | 25 |
| 1 | 1 | 1 | 1 | 1.E-04 | 2 | 1 | 1 | 10 |
| 1 | 1 | 1 | 1 | 1.E-04 | 2 | 1 | 1 | 1 |

***Methods – Additional Details on Statistical Analyses***

- Linear mixed effects models were fit using the restricted maximum likelihood approach and Kenward-Roger basis for degrees of freedom estimates via the lmerTest R package
- Significance of the main and interaction effects of the mixed effects models were determined using a Type III ANOVA with Satterthwaite's method.
- Post-hoc Tukey pairwise comparisons were made using the emmeans R package

***Methods – Sample Size Justification***

The sample population for this study was a subset of individuals with viable EMG data from an existing dataset (unpublished). To assess the appropriateness of the available sample population to test our hypothesis, we used the General Linear Mixed Model Power and Sample Size software (GLIMMPSE, version 3.0) to perform a power analysis to determine the necessary sample size to detect a 10% increase in total tibiofemoral contact force [5]. We informed the power analysis using our estimated total contact forces for the BASE conditions, our observed variability and correlation amongst the repeated measures (Task and Condition), and the hypothetical 10% increase in contact forces during the COG condition for all tasks. The GLIMMPSE settings/inputs file is available in an accompanying supplemental .json file.

***Results - CEINMS Results Quality Metrics***

$R^2$ between inverse dynamics and CEINMS sagittal plane hip, knee, and ankle moments were 97%±2%, 89%±4%, and 93%±5%, respectively. Normalized root mean squared errors between inverse dynamics and CEINMS sagittal plane hip, knee, and ankle moments were 2.7%±0.7%, 5%±1%, and 4%±2%, respectively. Across all trials in the final dataset, 85%±9% of EMG channels were deemed usable [6].

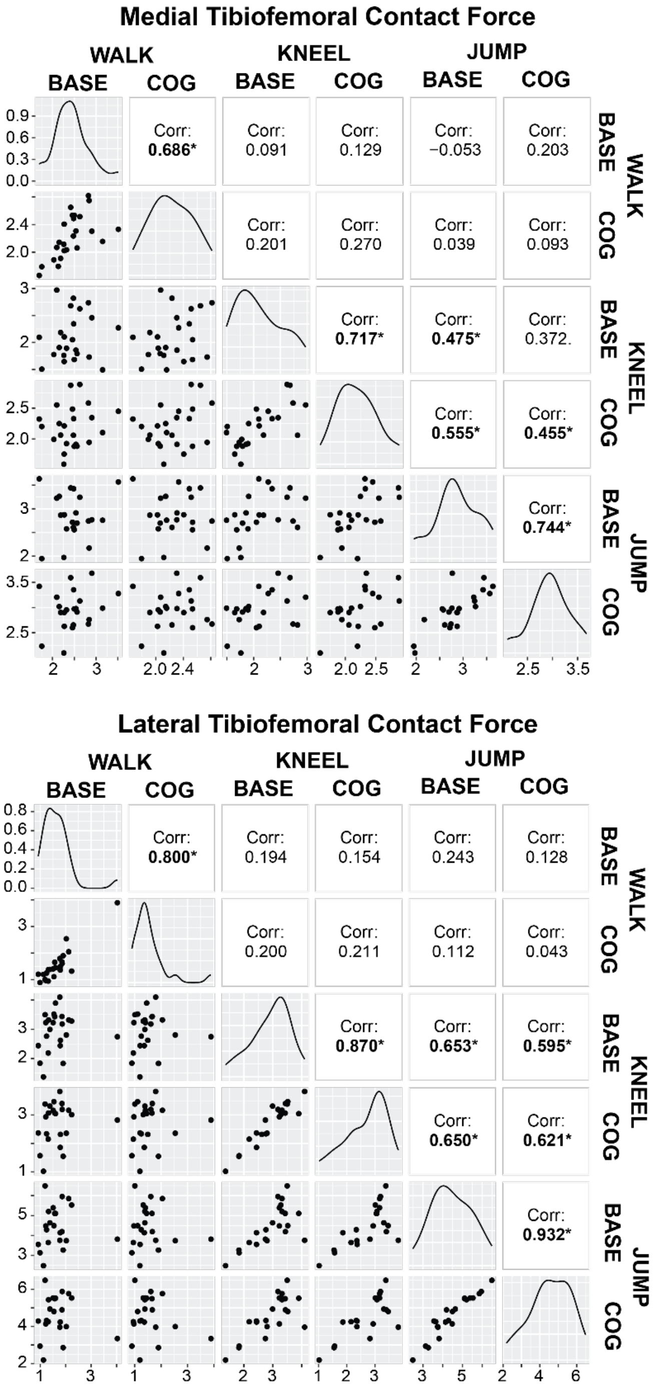


**Supplemental Figure 1**. Correlations for medial (top) and lateral (bottom) tibiofemoral contact forces across combinations of tasks and condition. Spearman correlation coefficients are reported in the upper right portion of the figure, with * indicating statistical significance. Lower left panels reflect the scatterplots of total tibiofemoral contact force with units of BW for the respective task-condition combination. The diagonal panels display the distribution of the variable.

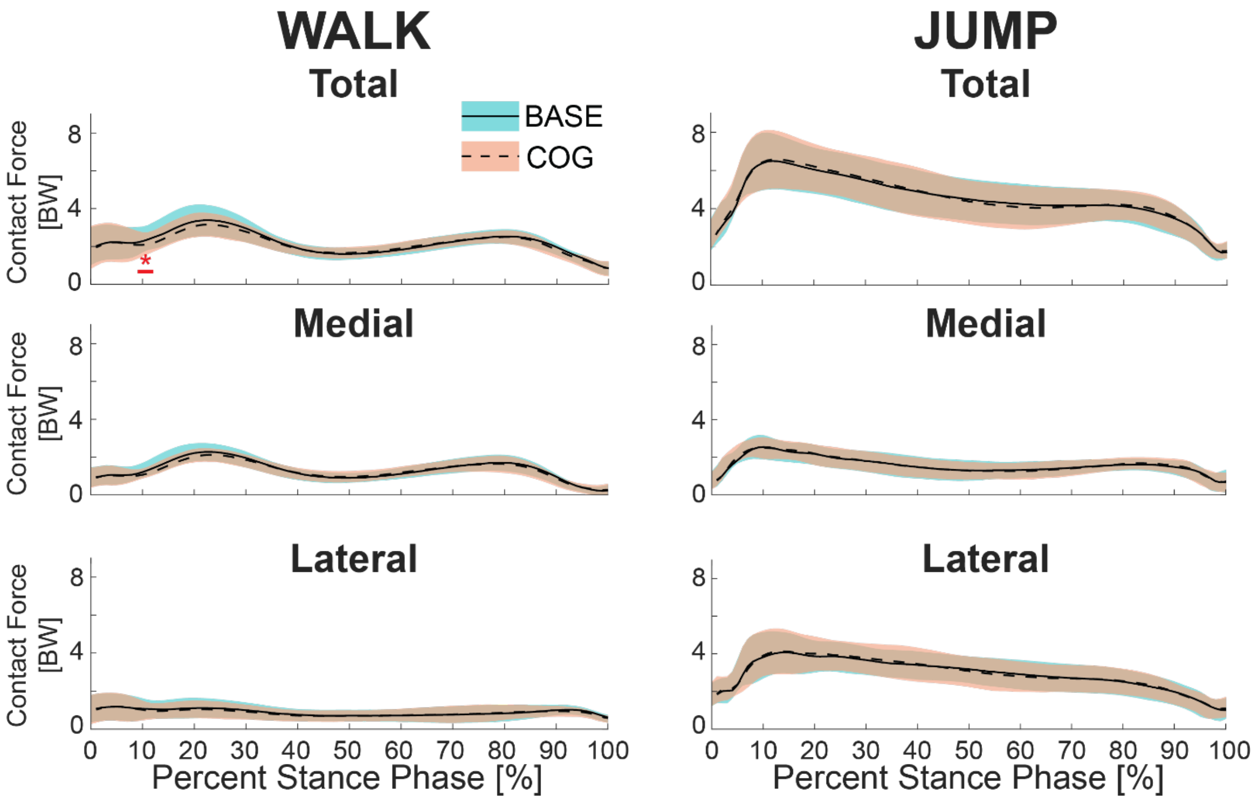


**Supplemental Figure 2**. Time series of total, medial, and lateral tibiofemoral contact forces throughout the WALK (left column) and JUMP (right column) tasks. Red lines with an asterisk along the x- axis indicate regions that are statistically different based on paired t-test SPM analyses.
